# Disruptions to schistosomiasis programmes due to COVID-19: an analysis of potential impact and mitigation strategies

**DOI:** 10.1101/2020.10.26.20219543

**Authors:** Klodeta Kura, Diepreye Ayabina, Jaspreet Toor, T. Deirdre Hollingsworth, Roy M. Anderson

**Affiliations:** London Centre for Neglected Tropical Disease Research, London, United Kingdom; Department of Infectious Disease Epidemiology, School of Public Health, Faculty of Medicine, St Mary’s Campus, Imperial College London, London, United Kingdom; MRC Centre for Global Infectious Disease Analysis; Big Data Institute, Li Ka Shing Centre for Health Information and Discovery, University of Oxford, Oxford OX3 7LF, UK; The DeWorm3 Project, The Natural History Museum of London, London, United Kingdom

**Keywords:** COVID-19, Elimination as a public health problem, Mass drug administration, Modelling, Schistosomiasis

## Abstract

**Background:** The 2030 goal for schistosomiasis is elimination as a public health problem (EPHP), with mass drug administration (MDA) of praziquantel to school-aged children (SAC) a central pillar of the strategy. However, due to COVID-19, many mass treatment campaigns for schistosomiasis have been halted with uncertain implications for the programmes.

**Method:** We use mathematical modelling to explore how postponement of MDA and various mitigation strategies affect achievement of the EPHP goal for *Schistosoma mansoni* and *S. haematobium*.

**Results:** In moderate and some high prevalence settings, the disruption may delay the goal by up to two years. In some high prevalence settings EPHP is not achievable with current strategies, and so the disruption will not impact this. Here, increasing SAC coverage and treating adults can achieve the goal.

The impact of MDA disruption and the appropriate mitigation strategy varies according to the baseline prevalence prior to treatment, the burden of infection in adults and stage of the programme.

**Conclusions:** Schistosomiasis MDA programmes in medium and high prevalence areas should restart as soon as is feasible, and mitigation strategies may be required in some settings.

## Introduction

Schistosomiasis is a parasitic disease affecting millions of people in several endemic regions. ^1^ Intestinal (caused by *Schistosoma mansoni* or *S. japonicum*) and urogenital (caused by *S. haematobium*) are the two most prevalent forms of human schistosomiasis. ^2^ At present, mass drug administration (MDA) of praziquantel to school-aged children (SAC; 5-14 years old) is the main method of reducing the burden of morbidity associated with this infection. ^3,4^ Control programmes additionally include recommending behaviour modification and improvements in sanitation to lower the intensity of transmission. ^5,6^

MDA is mostly targeted at school-aged children since age-intensity profiles are convex in shape with a peak in infection levels typically seen amongst SAC and teenagers. ^7,8^ Additionally, this age category can be reached through school-based treatment programmes which have been shown to be cost-effective in reaching these populations. ^9^ It should be noted that in some high-risk areas, treatment of adults is also recommended. ^10^

The 2030 World Health Organization (WHO) target for schistosomiasis is elimination as a public health problem (EPHP), achieved when the heavy-intensity prevalence in SAC reduces to less than 1%. ^11,12^ For *S. mansoni*, heavy-intensity infection is defined as having over 400 eggs per gram of feces and for *S. haematobium* is defined as having over 50 eggs per 10mL of urine. ^13^ Heavy-intensity infections can be diagnosed by using the Kato-Katz technique and urine filtration respectively. ^14–16^ Morbidity is thought to be associated most strongly with these heavy burdens, hence the target to reduce the number of these infections.

Previous mathematical modelling for schistosomiasis has shown that EPHP can be achieved in low (< 10%baseline prevalence among SAC) to moderate (10 - 50%baseline prevalence among SAC) transmission settings, but in certain high transmission settings (≥50%baseline prevalence among SAC) inclusion of adults in MDA programmes would be needed to achieve the EPHP goal.^2,11,17–19^

Due to the coronavirus 2019 (COVID-19) pandemic, the WHO has advised governments to postpone MDA for schistosomiasis (and other neglected tropical diseases). ^20^ It is likely that the MDA postponement will have a different impact in different transmission settings as the level of resurgence or bounce back varies across settings since it depends on the magnitude of the basic reproductive number, R_0_. Particularly, we expect the postponement to have a greater impact in high transmission settings, as they face the greatest risk since resurgence will be high in these areas. Although missed MDAs will certainly lead to resurgence in infection levels, the epidemiological impact of such postponement is not known.

The stage (how many rounds of MDA prior to the delay) and effectiveness of the programme (coverage and compliance) will clearly play a role in the resurgence or bounce back rate. Programmes in their early stages may return to pre-treatment endemicity levels faster, whereas, programmes further in which have managed to significantly reduce the intensity of transmission, will observe lower levels of resurgence, provided the transmission rate is not too high. However, in high transmission settings, programmes further in will have a risk of losing much of the long-term benefit of multiple rounds of MDA.

In this paper, we use mathematical models of parasite transmission and control by MDA to estimate the impact of temporarily delaying MDA on achieving the EPHP goal. Once MDA programmes resume, it will be of great importance for health workers and programme managers to have guidelines on mitigation strategies, in order to get back on track for achieving EPHP. We consider various stages of the programme at which the disruption occurs (early or late into the programme) and investigate the impact of mitigation strategies for *S. mansoni* and *S. haematobium*. For *S. mansoni* we also use two different age-intensity profiles, corresponding to low and high adult burdens of infection to determine if this would have an impact on missing MDA (**Figure 1**).

**Figure 1:**
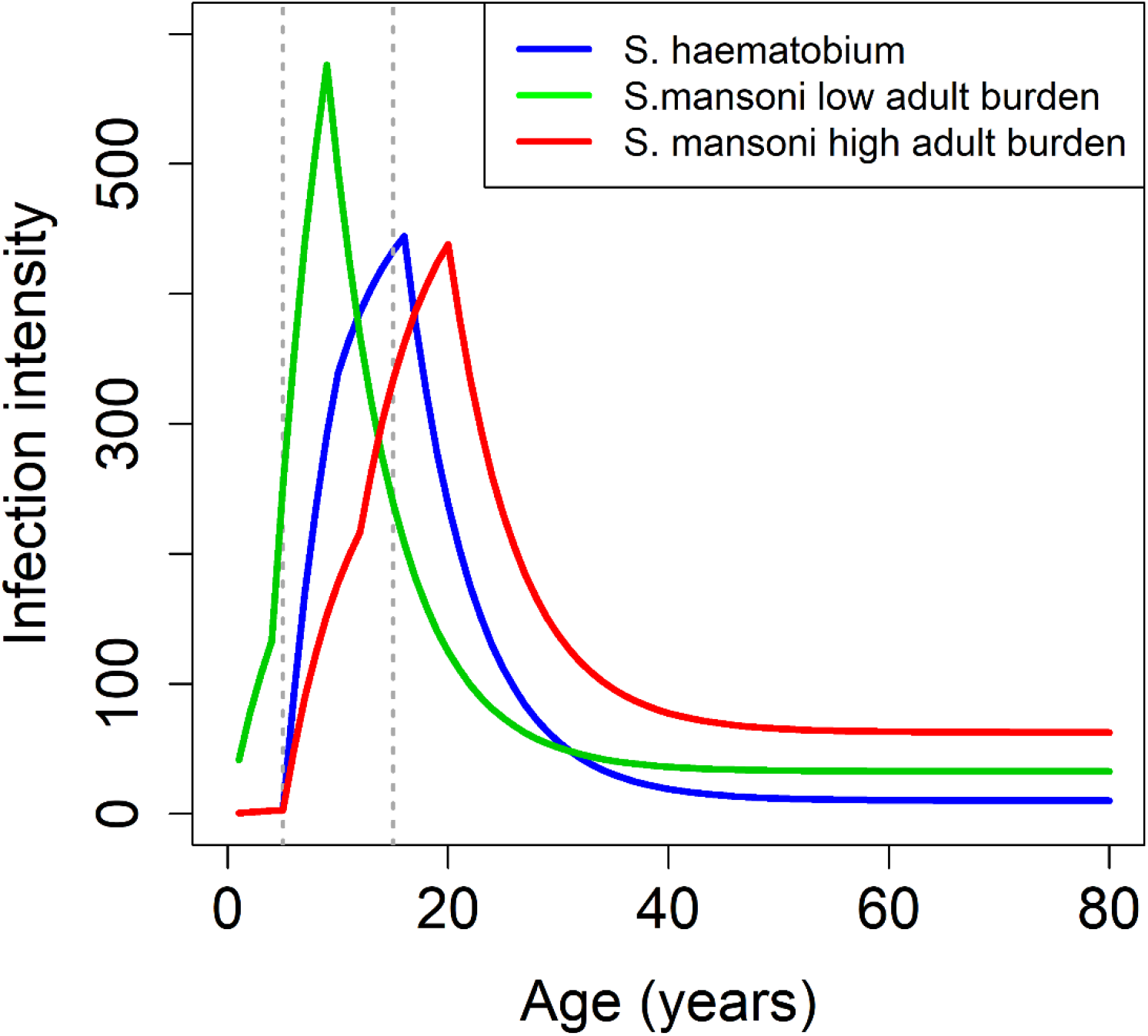
*S. mansoni* and *S. haematobium* age-intensity profiles of infection (eggs/10mL for *S. haematobium* and eggs/gram for *S. mansoni*, showing low and high burden of adult infection settings). ^17 21^

## Methods

### Transmission model

We adopt an age-structured deterministic model developed by Imperial College London (ICL). ^8,22^ This model has an individual based stochastic analogue ^8,22^ but the mean derived from this model is identical to the deterministic model prediction. The model incorporates treatment by MDA and is parameterised for *S. mansoni* and *S. haematobium* with previously published data and estimated parameter values derived from past epidemiological studies (supplementary **Table S1**). ^17,23^ Briefly, the model describes the dynamics of the adult worms in the human host population and a single reservoir of infectious material (infected snails-are short lived). ^22^ This model assumes a negative binomial distribution of parasites per host with a fixed aggregation parameter, k, density-dependent fecundity, and assumes monogamous sexual reproduction among worms. The egg contribution to the reservoir depends on the age-specific contact rate for each individual in the population.

The numerical simulations were run for a single community with a population size set at 1000, assuming no migration. In our simulations, treatment is delivered at random at each round i.e. no systematic non-adherers and no non-access to treatment individuals. Acquired immunity is not taken into consideration. ^24^ To simulate moderate and high baseline prevalence settings (for low and high adult burden of infection), the intrinsic intensity of transmission, i.e. basic reproductive number (R_0_), is varied (higher prevalence settings corresponding to higher R_0_ values).

### Scenarios and mitigation strategies

In our investigation, we consider moderate (10 − 50%baseline prevalence among SAC) and high (≥50%baseline prevalence among SAC) prevalence settings prior to MDA. ^2,11^ In addition to this, for *S. mansoni* we use two different age intensity profiles (low and high adult burden of infection) to determine whether this would differentially influence the impact of missing MDA. In the model, we implement MDA annually at a 75% coverage level of SAC only. ^11^ We simulate one MDA round being missed either early or late (second or sixth round of MDA, respectively) into the programme. For all of our scenarios, we determine the time taken to achieve EPHP. After a missed round of MDA, we consider three mitigation strategies (**Figure 2**): (i) return with annual 75% coverage level of SAC-only, (ii) return with annual 85% coverage level of SAC only and (iii) return with one-year community-wide coverage (85% SAC +40% adults) followed by 75% coverage of SAC-only in the years following this.

**Figure 2:**
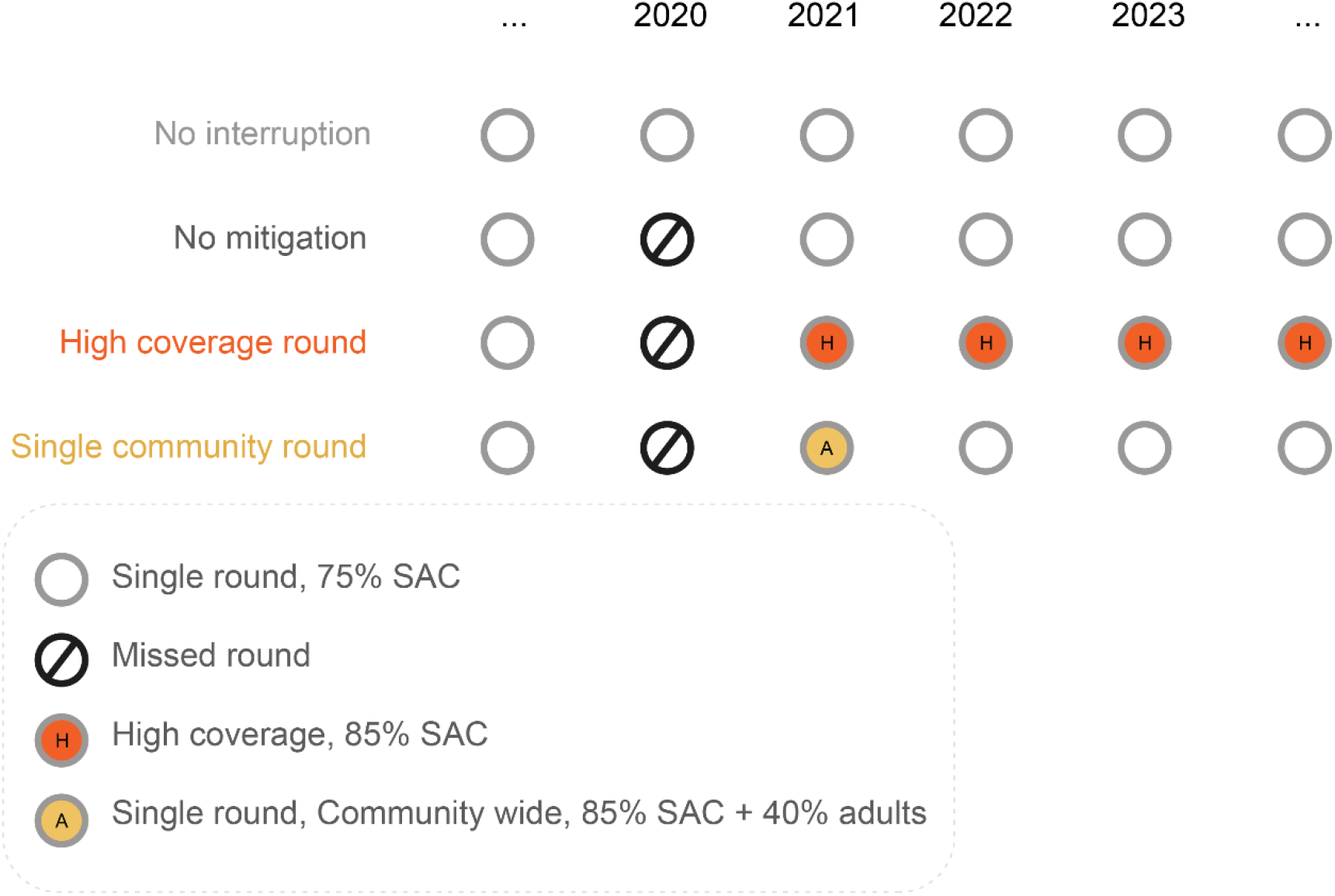
Visual representation of the scenarios and mitigation strategies analysed.

For each transmission setting and age profile (**Figure 1**), we simulate the impact of the different control strategies over a period of 15 years. For each point in time, we determine the prevalence of heavy-intensity infections (eggs per gram, epg ≥ 400 for *S. mansoni* and over 50 eggs per 10mL of urine for *S. haematobium*) in SAC to investigate whether the EPHP goal has been achieved.

## Results

We present results for the effect of MDA postponement due to COVID-19 and the impact of mitigation strategies in going back on track to achieving EPHP by 2030. The results are presented for *Schistosoma mansoni* and *S. haematobium* by considering the scenarios and mitigation strategies described in **Figure 2**.

## Results for *Schistosoma mansoni*

For moderate transmission settings with a low or high adult burden of infection, missing the second round of MDA (refer to **Table 1**), requires an additional year of intervention to achieve EPHP, regardless of the mitigation strategy. It should be noted that in lower moderate transmission settings (i.e. just above 10% SAC prevalence), with 75% coverage and random compliance the EPHP goal is achieved after one round of MDA so there is no delay to the goal when the second MDA is missed. Missing the sixth round of MDA does not have any impact on the time required to achieve the EPHP goal as the goal has already been reached prior to the sixth round (refer to **Table 2**).

**Table 1:** Years of MDA to achieve EPHP (≤1% heavy-intensity prevalence in SAC) for *S. mansoni*. The second round of MDA is missed. NA: not achievable by 2030 (for baseline higher than 59% in SAC). Results are shown for low and high adult burden of infection settings using the Imperial College London deterministic model.

| Prevalence in SAC | Moderate (10 – 50%) | High ($\geq 50\%$) |
| --- | --- | --- |
| **Time to EPHP if no postponement to annual 75% SAC MDA** | Low adult burden: 1-3 years<br>High adult burden: 1-3 years | Low adult burden: 3-8 years<br>High adult burden: 3 – NA years |
| **Delay to EPHP if 2<sup>nd</sup> MDA missed + return with 75% SAC** | Low adult burden: 0- 1 years<br>High adult burden: 0 – 1 years | Low adult burden: 1-2 years<br>High adult burden: 1 – NA years |
| **Delay to EPHP if 2<sup>nd</sup> MDA missed + return with 85% SAC** | Low adult burden: 0 - 1 years<br>High adult burden: 0 – 1 years | Low adult burden: 1 - 0 years<br>High adult burden: 1 - NA years |
| **Delay to EPHP if 2<sup>nd</sup> MDA missed + return with 1 community-wide MDA (85% SAC + 40% adults) followed by 75% SAC** | Low adult burden: 0 - 1 years<br>High adult burden: 0 -1 years | Low adult burden: 1-1 years<br>High adult burden: 1 - NA years |

**Table 2:** Years of MDA to achieve EPHP (≤1% heavy-intensity prevalence in SAC) for *S. mansoni*. The sixth round of MDA is missed. NA: not achievable by 2030. Results are shown for low and high adult burden of infection settings using the Imperial College London deterministic model

| Prevalence in SAC | Moderate (10 – 50%) | High ($\geq 50\%$) |
| --- | --- | --- |
| **Time to EPHP if no postponement to annual 75% SAC MDA** | Low adult burden: 1-3 years<br>High adult burden: 1-3 years | Low adult burden: 3-8 years<br>High adult burden: 3 – NA years |
| **Delay to EPHP if 6<sup>th</sup> MDA missed + return with 75% SAC** | Low adult burden: 0 years<br>High adult burden: 0 years | Low adult burden: 0- 2 years<br>High adult burden: 0 – NA years |
| **Delay to EPHP if 6<sup>th</sup> MDA missed + return with 85% SAC** | Low adult burden: 0 years<br>High adult burden: 0 years | Low adult burden: 0 - 2 years<br>High adult burden: 0 - NA years |
| **Delay to EPHP if 6<sup>th</sup> MDA missed + return with 1 community-wide MDA (85% SAC + 40% adults) followed by 75% SAC** | Low adult burden: 0 years<br>High adult burden: 0 years | Low adult burden: 0 – 2 years<br>High adult burden: 0 – NA years |

For high transmission settings with a low adult burden of infection, if the programme is reintroduced at the previous 75% SAC-only coverage, then it is predicted that up to two years of delay will result in reaching EPHP (**Tables 1, 2** and **Figure 3**), regardless of the time MDA is missed. Increasing coverage level to 85% of SAC (or having one round of community-wide MDA), requires up to one additional year if the second round of MDA is missed. From **Figure 3**, during the postponement of MDA there is an increase in the heavy – intensity infection (illustrated by the black, red, and yellow lines). As a result of this there is an increase in prevalence of heavy intensity (related to increased morbidity), illustrated by the green area. Hence, this is an additional burden of infection which would not have happened if the treatment programme had gone as planned.

**Figure 3:**
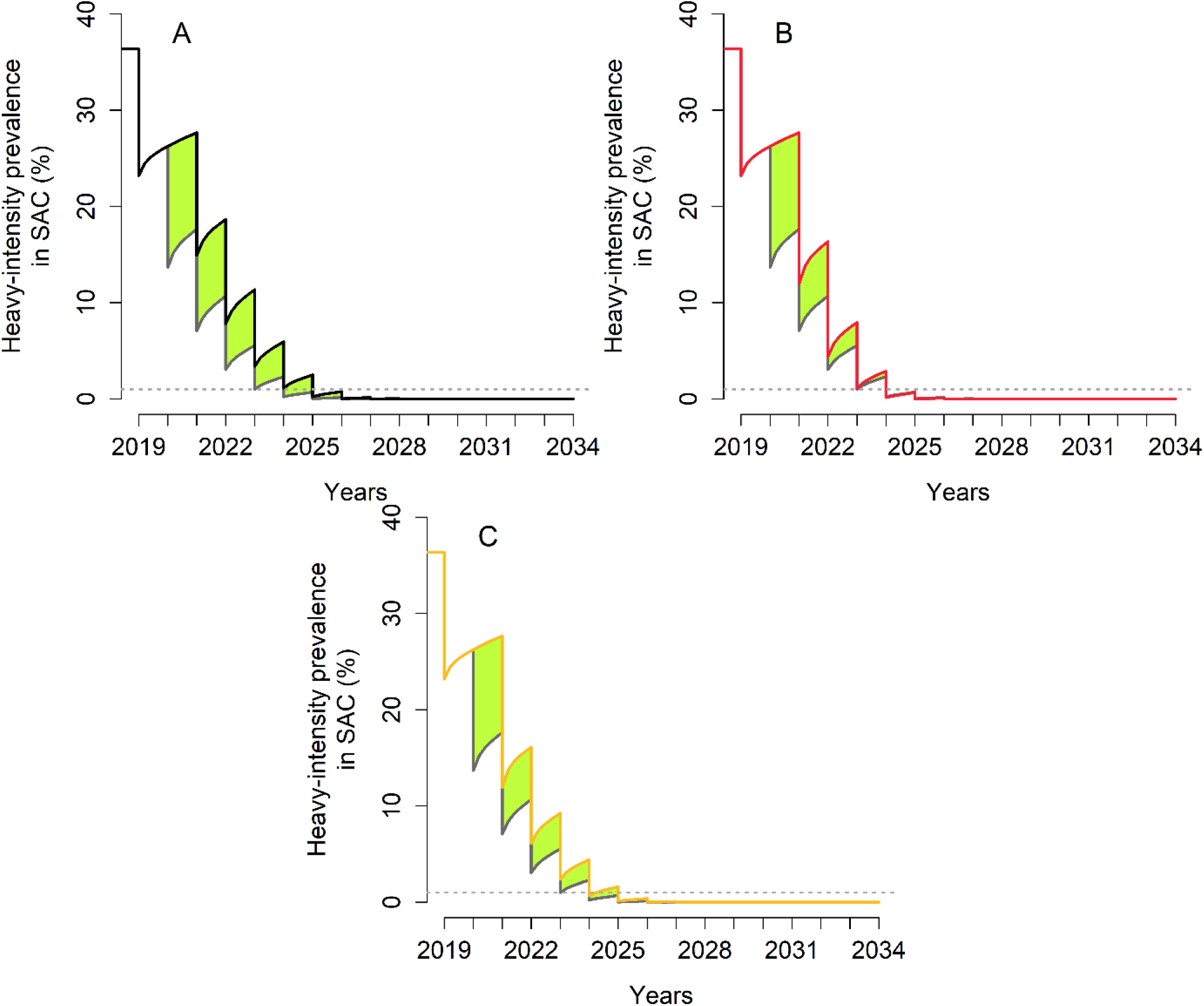
Heavy-intensity prevalence in SAC for *S. mansoni* in high transmission settings with a low adult burden of infection. The second round of MDA is missed. The grey line gives the prevalence of heavy infection if the treatment had gone ahead as planned. (**A**) the programme is restarted by treating 75% of SAC (black line). (**B**) the programme is restarted by treating 85% of SAC (red line). (**C**) the programme is restarted with one community-wide MDA (85% SAC + 40% adults) followed by 75% SAC (yellow line). The green area shows the increased level of infection in the community.

For high transmission settings with a high adult burden, the outcome depends on the baseline SAC prevalence. For baseline SAC prevalence below 59%, missing the second round of MDA, a one-year delay in achieving EPHP is predicted. This holds for any mitigation strategy considered. However, for baseline SAC prevalence above 59%, EPHP is not achieved by 2030 regardless of the mitigation strategy described in **Figure 2** (refer to **Table 1, Table 2** and **Figure 4A, Figure 4B, Figure 4C**). This is because MDA of SAC only is having a small impact on reducing transmission. To achieve EPHP within a shorter time frame, higher coverage of SAC and treating adults would be needed for this setting. These coverage levels can be determined by collecting SAC and adult data once programmes resume. ^17^ In **Figure 4D**, it is shown that once the MDA programme resumes, increasing the SAC coverage to 85% and including 40% of adults for every MDA round, can achieve the goal by 2030.

**Figure 4:**
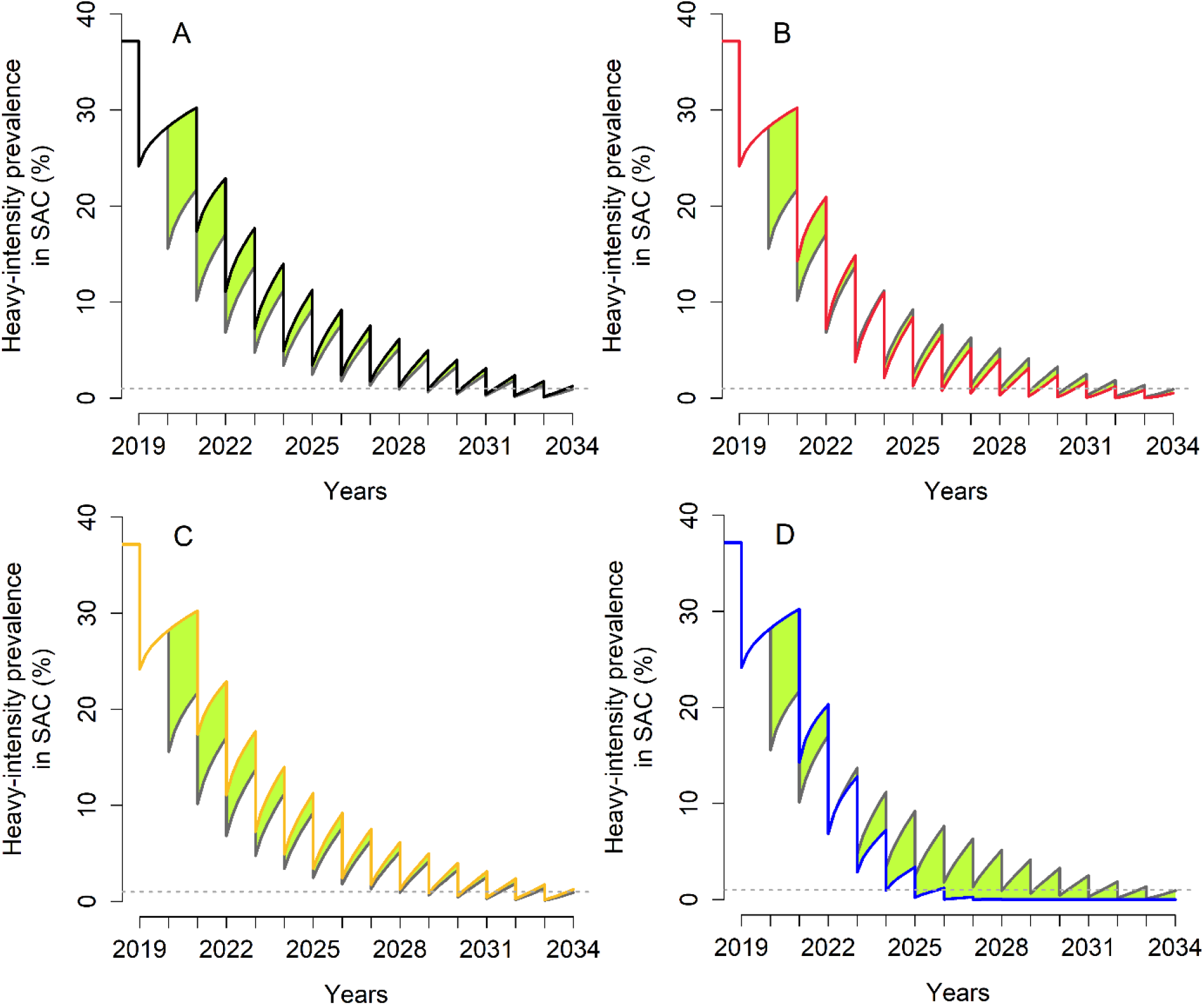
Heavy-intensity prevalence in SAC for *S. mansoni* in high transmission settings with a high adult burden of infection. The second round of MDA is missed. The grey line gives the prevalence of heavy infection if the treatment had gone ahead as planned. (**A**) the programme is restarted by treating 75% of SAC (black line). (**B**) the programme is restarted by treating 85% of SAC (red line). (**C**) the programme is restarted with one community-wide MDA (85% SAC + 40% adults) followed by 75% SAC (yellow line). (**D**) the programme is restarted by treating 85% of SAC and 40% of adults (blue line). The green area shows the increased level of infection in the community.

If the sixth round of MDA is missed and the baseline SAC prevalence is below 59%, no additional year of intervention is required, regardless of the mitigation strategy. However, for baseline SAC prevalence above this threshold, EPHP is not achieved by 2030 with any of the mitigation strategies considered in **Figure 2**. Similarly, as when the second round of MDA is postponed, increasing the SAC coverage to 85% and treating 40% of adults in every round after the programme resumes, can achieve the EPHP goal by 2030.

Our simulations show that missing the second round of MDA for baseline SAC prevalence of 30% (moderate transmission setting), may take from four to ten years for SAC prevalence to catch up to the state where no MDA rounds are missed (depending on the scenario and adult burden of infection, refer to supplementary **Table S2** and **Table S3**). Missing the sixth round of MDA does not have any impact on the time required to achieve the EPHP goal but it might take up to five years for the SAC prevalence to catch up to what would have been achieved without missing MDA rounds.

For a baseline SAC prevalence of 70% (high transmission setting) with a low adult burden, it may take from five to 12 years for the SAC prevalence to catch up (supplementary **Table S2**). For the high transmission setting with a high adult burden, it is predicted that it may take more than three years for the SAC prevalence to get back to the level under no missed rounds (depending on the scenario, refer to supplementary **Table S3**).

## Results for *S. haematobium*

Here, results of the impact of COVID-19 on *S. haematobium*, and the effect of mitigation strategies on getting the programmes back on track when MDA resumes are presented.

For moderate transmission settings with no postponement of MDA, it takes up to two years for the elimination as a public health problem to be achieved (**Table 3** and **Table 4**). For lower moderate prevalence settings (i.e. just above 10% SAC prevalence), the heavy-intensity prevalence in SAC may be less than 1% before the start of the treatment. Therefore, EPHP goal is met without any MDA intervention. Missing the second round of MDA (**Table 3**), will require up to one additional year to achieve the goal, regardless of the mitigation strategy. However, missing the sixth round of MDA does not have any effect on the goal because it has been achieved prior to the missed MDA (**Table 4**).

**Table 3:** Years of MDA to achieve EPHP (≤1% heavy-intensity prevalence in SAC) for *S. haematobium*. The second round of MDA is missed.

| Prevalence in SAC | Moderate (10 – 50%) | High ($\geq 50\%$) |
| --- | --- | --- |
| Time to EPHP if no postponement to annual 75% SAC MDA | 0 – 2 years | 2-9 years |
| Delay to EPHP if 2<sup>nd</sup> MDA missed + return with 75% SAC | 0 – 1 years | 1 year |
| Delay to EPHP if 2<sup>nd</sup> MDA missed + return with 85% SAC | 0 – 1 years | 1 - 0 years |
| Delay to EPHP if 2<sup>nd</sup> MDA missed + return with 1 community-wide MDA (85% SAC + 40% adults) followed by 75% SAC | 0 – 1 years | 1-0 years |

**Table 4:**
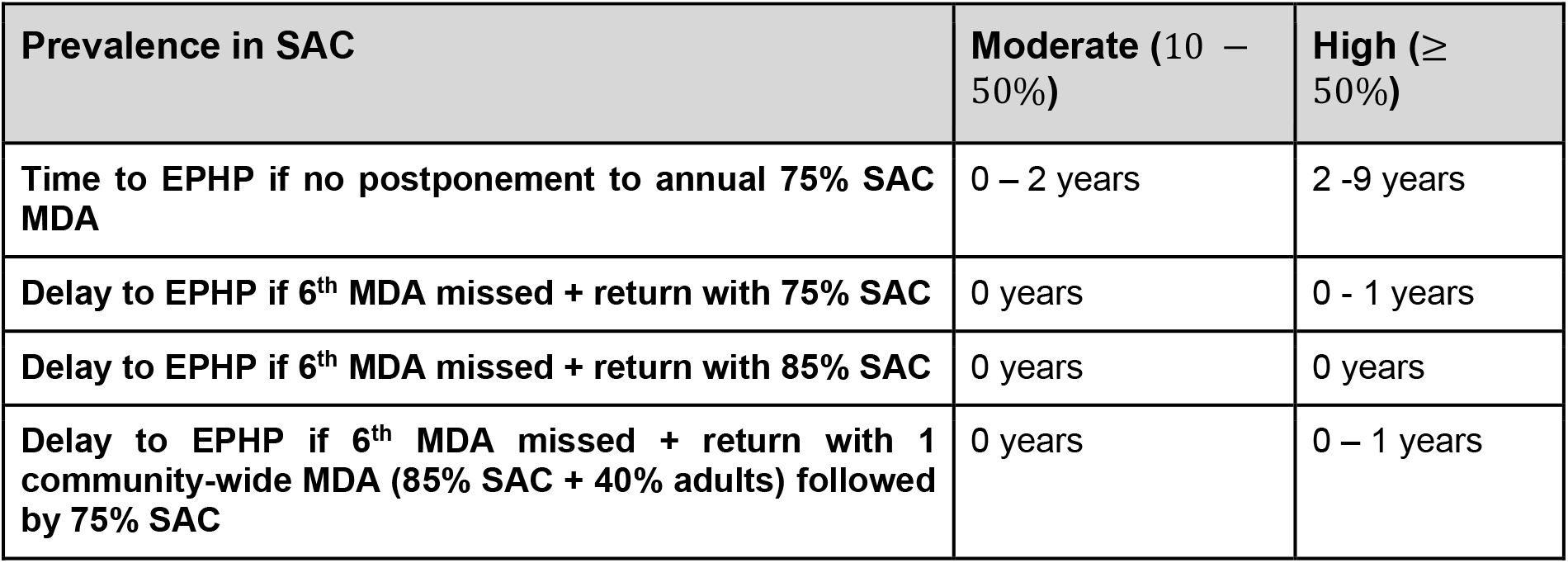
Years of MDA to achieve EPHP (≤1% heavy-intensity prevalence in SAC) for *S. haematobium*. The sixth round of MDA is missed.

| Prevalence in SAC | Moderate (10 – 50%) | High ($\geq 50\%$) |
| --- | --- | --- |
| Time to EPHP if no postponement to annual 75% SAC MDA | 0 – 2 years | 2 -9 years |
| Delay to EPHP if 6<sup>th</sup> MDA missed + return with 75% SAC | 0 years | 0 - 1 years |
| Delay to EPHP if 6<sup>th</sup> MDA missed + return with 85% SAC | 0 years | 0 years |
| Delay to EPHP if 6<sup>th</sup> MDA missed + return with 1 community-wide MDA (85% SAC + 40% adults) followed by 75% SAC | 0 years | 0 – 1 years |

For high transmission settings, depending on the baseline SAC prevalence, it takes two to nine years to achieve EPHP (no delay to MDA treatment). For scenarios where it takes two years to EPHP, missing the second round of MDA will require one additional year of intervention, regardless of the mitigation strategies. For scenarios where it takes longer than two years to achieve EPHP, a one year delay is also expected when the programme is reintroduced at the previous coverage level after missing the second round of MDA (refer to **Table 3** and **Figure 5**). However, increasing the coverage level to 85% SAC (or having one round of community-wide treatment), does not require the additional year of MDA. Missing the sixth round of MDA has a smaller effect on the time to achieve the goal. Increasing the coverage level to 85% SAC only, does not require any additional years of treatment (**Table 4**).

**Figure 5:**
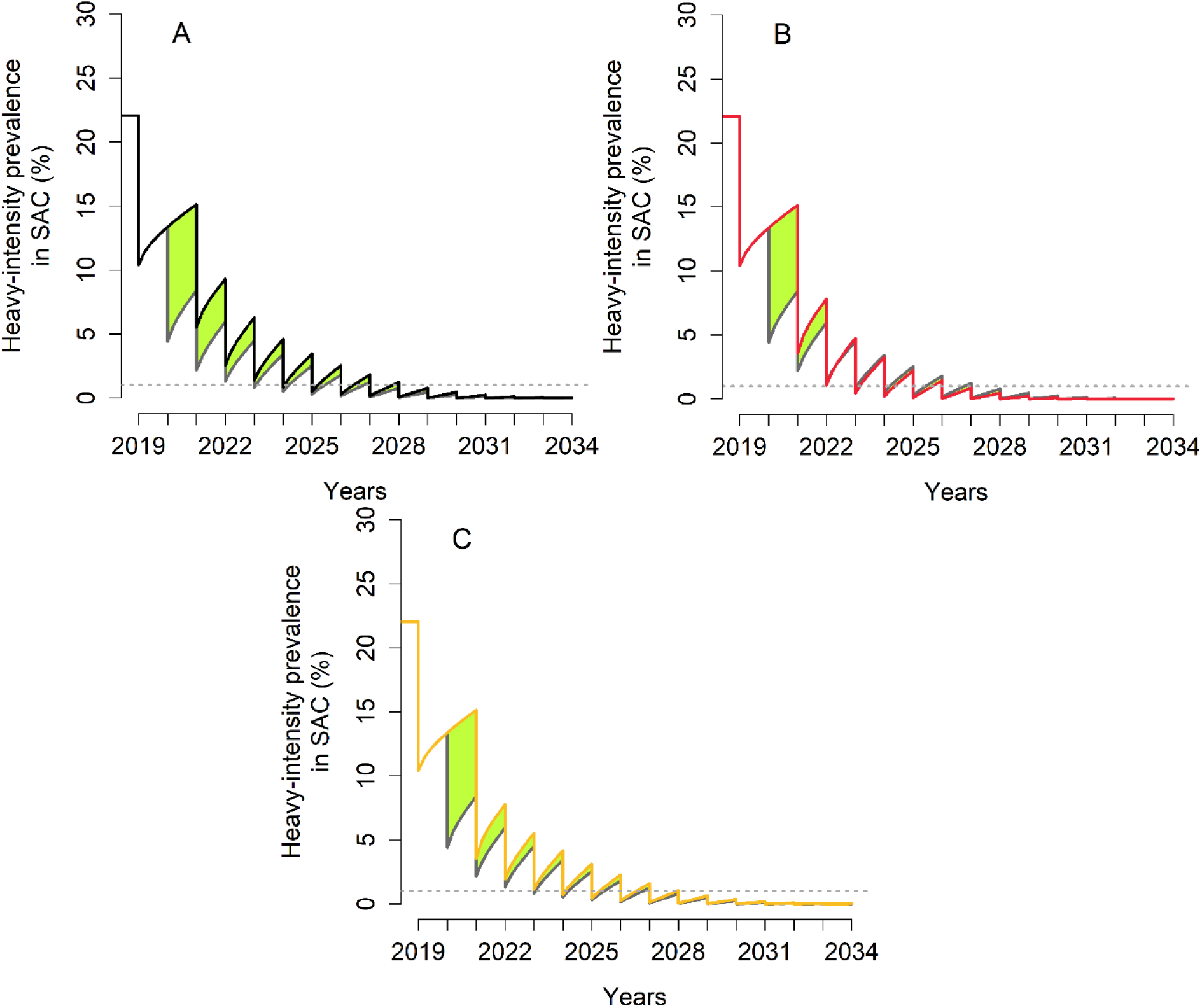
Heavy-intensity prevalence in SAC for *S. haematobium*. The second round of MDA is missed. The grey line gives the prevalence of heavy infection if the treatment had gone ahead as planned. (**A**) the programme is restarted by treating 75% of SAC (black line). (**B**) the programme is restarted by treating 85% of SAC (red line). (**C**) the programme is restarted with one community-wide MDA (85% SAC + 40% adults) followed by 75% SAC (yellow line). The green area shows the increased level of infection in the community.

## Discussion

We have presented analyses of the impact of delaying MDA due to COVID-19 spread and considered various mitigation strategies to get the programme back on track for achieving EPHP by 2030 for both *S. mansoni* and *S. haematobium*. We assumed that MDA will be delayed for one year, either early or late into the programme (second or sixth round of treatment). Once the programme resumes, the delay in achieving EPHP is calculated for various mitigation strategies.

For *S. mansoni*, our analyses suggest that in moderate transmission settings treating 75% of SAC annually can achieve EPHP within three years. This holds for low and high adult burden of infection. Missing the second round of MDA can delay EPHP by up to one year, whereas missing the sixth round of MDA does not have any effect on EPHP. This is because the goal is achieved before the delay occurs and hence it takes longer to resurge back to pre-MDA levels. In high transmission settings, the results depend on the baseline prevalence and the burden of infection in adults. For low adult burden of infection, treating 75% of SAC annually can achieve EPHP within eight years of MDA treatment. Missing one round of MDA, delays the goal by up to two years. For high adult burden of infection, we might not achieve the EPHP goal, regardless of the postponement. If the baseline SAC prevalence rises above 59%, and the second or the sixth round of MDA is missed, an increase in SAC coverage and inclusion of adults is necessary to achieve EPHP by 2030.

We acknowledge that due to limited praziquantel supplies (donations), ^25^ including adults in treatment may not be feasible in all areas. Hence, it is important that surveys are conducted to collect SAC and adult data to determine the optimal coverage levels and whether adult treatment is required. ^23^ This will then allow for community-wide treatment to be prioritised as necessary in high transmission settings where there is a high adult burden of infection.

For *S. haematobium*, it is predicted that in moderate transmission settings, missing the second round of MDA delays EPHP by up to one year. However, missing the sixth round of MDA does not affect the goal. In high transmission settings, EPHP can be delayed by up to one year, regardless of the time of the delay. Annual 75% SAC only treatment is sufficient for achieving EPHP by 2030, even when MDA is postponed for one year.

Overall, postponing MDA for one-year results in a delay of up to two years for achieving EPHP. The impact of missing MDA depends on the baseline prevalence prior to treatment, the burden of infection in adults and the time at which we miss MDA (early or late into the programme).

However, care should be taken when deciding to stop MDA after EPHP has been achieved as there will be a risk of resurgence/bounce back. This is because the overall prevalence might still be high, so that infection persists, despite the heavy-intensity prevalence in SAC being reduced to less than 1%. ^26^ Additionally, it is predicted that it takes longer for the SAC prevalence to catch-up to what would have been achieved by full MDA rounds, than it takes for the heavy-intensity prevalence.

In this study, we assume that control programmes will return to their pre-COVID-19 effectiveness within one year, but this might not be feasible for various reasons. Training programmes may have been disrupted by the COVID-19 and health workers might have been re-deployed to other tasks. Another important factor is that schools may not open when the programme restarts or parents may decide not to send their children back to school. As the MDA programme is mainly focused in SAC, this will have a major impact on the mitigation strategies. We also need to take into consideration that stocks of praziquantel in government warehouses may have past their expiry dates, or that praziquantel production and supply chains of MDA treatment may be disrupted due to travel restrictions. Thus, it might take some time to achieve the desired coverage once the programme restarts. As a result, postponing programmes for longer or returning with reduced effectiveness will mean that we might ultimately be facing longer delays in achieving the EPHP goal.

When considering a longer postponement of MDA, of 18 months, analyses suggest that the EPHP goal could be delayed by an extra six months, depending on the transmission setting and adult burden of infection. Hence, the longer the delay, the longer it will take programmes to achieve EPHP. Mitigation strategies upon resumption will be increasingly important in areas where programmes are delayed for longer.

It is important to note one important caveat on the predictions made in this study. It has been assumed that for a fixed MDA coverage level, treatment is at random in the population. This may not be the case, as persistent non-adherers to treatment (due to many different factors) are an important feature of most MDA based control programmes. If this is the case for treating schistosome infections, our predictions may err on the side of being too optimistic.

The model-based predictions can be tested once the MDA programme is resumed as we expect to see a large increase in prevalence of infection after a long period of no intervention, particularly in high transmission settings. Data collection on SAC and adults needs to be done at the start of the resumed intervention. The ongoing Geshiyaro project can address this. ^27^

## Conclusions

In this study we show that postponement of rounds of MDA due to the COVID-19 pandemic will lead to an increase in *S. mansoni* and *S. haematobium* infection. As a result, more resources will be needed to reach the 2030 goal of EPHP once the full MDA programmes restart. The transmission setting, duration of the delay in delivering MDA, stage of programme, age-intensity profile will all have an impact on achieving WHO goals for control of both morbidity and transmission. Mitigation strategies can help in accelerating progress towards EPHP by 2030. In some high transmission settings, EPHP may not be reached regardless of the length of the delay and hence, upon resumption it is important that surveys are done to collect SAC and adult infection data in order to determine the desired coverage levels for MDA to reach the defined control objectives. We hope this study will provide health workers with important quantitative tools to assess what mitigation strategies are best applied in given epidemiological settings.

## Data Availability

All data generated or analyzed during this study are included in this published article.

## Authors’ contributions

KK, DA, JT, TDH and RMA conceived and designed the study. KK carried out the analysis, interpreted the outputs and drafted the manuscript. DA, JT, TDH and RMA reviewed and edited the manuscript. All authors read and approved the final manuscript.

## Acknowledgements

We thank Simon Brooker, Amadou Garba Djirmay and Andreia Vasconcelos for helpful comments and discussions on the study.

## Ethical approval

Not required.

