## Supplementary Information for "Disruptions to schistosomiasis programmes due to COVID-19: an analysis of potential impact and mitigation strategies"

**Table S1:** Parameter values used for *Schistosoma mansoni* and *S. haematobium*.

| **Parameter** | **Value** | **Source** |
| --- | --- | --- |
| Fecundity | *S. mansoni*: 0.34 eggs/female/sample  *S. haematobium*: 0.3 | ^1–4^ |
| Egg distribution within the individual | *S. mansoni*: 0.87  *S. haematobium*: 0.5 | ^1,2,4^ |
| Aggregation parameter | 0.24 | ^1,5–8^ |
| Density dependent fecundity | *S. mansoni*: 0.0007/female worm  *S. haematobium*: 0.0006/female worm | ^1,5,9^ |
| Worm lifespan | *S. mansoni*: 5.7 years  *S. haematobium*: 4 years | ^1,2,10^ |
| Drug efficacy | *S. mansoni*: 86.3%  *S. haematobium*: 94% | ^1,11^ |
| *Mansoni* low adult burden setting: Age specific contact rates for 0-4, 5-9, 10-15, 16+ years old | 0.01, 1.2, 1, 0.02 | ^9,12^ |
| *Mansoni* high adult burden setting: Age specific contact rates for 0-4, 5-11, 12-19, 20+ years old | 0.01, 0.61, 1, 0.12 | ^9,12^ |
| *Haematobium* age specific contact rates for 0-4, 5-9, 10+ years old | 0.3, 1, 0.02 | ^1^ |
| Prevalence of infection | Percentage of population having > 0 eggs per gram [epg] (*S. mansoni*) or > 0 eggs/10ml (*S. haematobium*) | - |
| Heavy-intensity infection prevalence | Percentage of population having ≥ 400 epg (*S. mansoni*) or ≥ 50 eggs/10ml (*S. haematobium*) | ^13,14^ |
| Human demography | Based on Uganda’s demographic profile | ^15,16^ |

**Table S2: Low adult burden of infection for *S. mansoni*.** Years of MDA required for the SAC prevalence to catch-up after the programme is resumed (the second or the sixth round of MDA is missed for one year).

| **Prevalence in SAC prior to treatment** | **Annual 75% SAC MDA is resumed** | **Annual 85% SAC MDA is resumed** | **One round of community-wide MDA is delivered, before returning to annual 75% SAC MDA** |
| --- | --- | --- | --- |
| Moderate (10-50%)    Baseline prevalence in SAC: 30% | **Miss the 2^nd^ round:** SAC prevalence catches up after 6 years.  **Miss the 6^th^ round:** SAC prevalence catches up after 3 years. | **Miss the 2^nd^ round:** SAC prevalence catches up after 5 years.  **Miss the 6^th^ round:** SAC prevalence catches up after 2 years. | **Miss the 2^nd^ round:** SAC prevalence catches up after 6 years.  **Miss the 6^th^ round:** SAC prevalence catches up after 2 years. |
| High (≥50%)    Baseline prevalence in SAC: 70% | **Miss the 2^nd^ round:** SAC prevalence catches up after 12 years.  **Miss the 6^th^ round:** SAC prevalence catches up after 8 years. | **Miss the 2^nd^ round:** SAC prevalence catches up after 5 years.  **Miss the 6^th^ round:** SAC prevalence catches up after 5 years. | **Miss the 2^nd^ round:** SAC prevalence catches up after 9 years.  **Miss the 6^th^ round:** SAC prevalence catches up after 6 years. |

**Table S3: High adult burden of infection for *S. mansoni*.** Years of MDA required for the SAC prevalence to catch-up after the programme is resumed (the second or the sixth round of MDA is missed for one year).

| **Prevalence in SAC prior to treatment** | **Annual 75% SAC MDA is resumed** | **Annual 85% SAC MDA is resumed** | **One round of community-wide MDA is delivered, before returning to annual 75% SAC MDA** |
| --- | --- | --- | --- |
| Moderate (10-50%)    Baseline prevalence in SAC: 30% | **Miss the 2^nd^ round:** SAC prevalence catches up after 10 years.  **Miss the 6^th^ round:** SAC prevalence catches up after 5 years. | **Miss the 2^nd^ round:** SAC prevalence catches up after 4 years.  **Miss the 6^th^ round:** SAC prevalence catches up after 3 years. | **Miss the 2^nd^ round:** SAC prevalence catches up after 4 years.  **Miss the 6^th^ round:** SAC prevalence catches up after 3 years. |
| High (≥50%)    Baseline prevalence in SAC: 70% | **Miss the 2^nd^ round:** SAC prevalence does not catch up  **Miss the 6^th^ round:** SAC prevalence does not catch up. | **Miss the 2^nd^ round:** SAC prevalence catches up after 3 years.  **Miss the 6^th^ round:** SAC prevalence catches up after 3 years. | **Miss the 2^nd^ round:** SAC prevalence catches up after 5 years.  **Miss the 6^th^ round:** SAC prevalence catches up after 3 years. |

To increase the modelling impact to policy and decision makers, we summarize in **Table S4** the five modelling principles, as described in, ^17^ and how they are achieved in the manuscript.

**Table S4:** The Policy-Relevant Items for Reporting Models in Epidemiology of Neglected Tropical Diseases (PRIME-NTD) .^17^

| **Principle** | **What has been done to satisfy the principle?** | **Where in the manuscript is this described?** |
| --- | --- | --- |
| **Stakeholder engagement** | Work has been presented at the following WHO webinars: (i) Neglected Tropical Diseases and COVID-19: Impact on Programme Implementation; and (ii) A Research Agenda for NTD Programmes Affected by the COVID-19 Pandemic. | - |
| **Complete model documentation** | Transmission model and mitigation strategies are described in the manuscript. | Methods section |
| **Complete description of data used** | Data and parameters used are described in the manuscript. | Figure 1 and Table S1 |
| **Communicating uncertainty** | We have considered two age-intensity profiles for S. mansoni and different stages of the programme for MDA interruption. | Methods and Results sections |
| **Testable model outcomes** | The model outcomes can be tested by the ongoing Geshiyaro project and by collecting data once programmes resume. | Discussion section |

13.	WHO Expert Committee on the Control of Schistosomiasis. *Prevention and Control of Schistosomiasis and Soil-Transmitted Helminthiasis : Report of a WHO Expert Committee.* World Health Organization; 2002.

14.	WHO | Schistosomiasis: progress report 2001–2011, strategic plan 2012–2020. *WHO*. Published online 2017. Accessed April 30, 2019. https://www.who.int/neglected_diseases/resources/9789241503174/en/

15.	Anderson R, Truscott J, Hollingsworth TD. The coverage and frequency of mass drug administration required to eliminate persistent transmission of soil-transmitted helminths. *Philos Trans R Soc B Biol Sci*. Published online 2014. doi:10.1098/rstb.2013.0435

16.	Pullan RL, Kabatereine NB, Quinnell RJ, Brooker S. Spatial and genetic epidemiology of hookworm in a rural community in Uganda. *PLoS Negl Trop Dis*. Published online 2010. doi:10.1371/journal.pntd.0000713
